# iPlayer: an open-source tool for time-windowed video annotation of human physical activity and behaviour

**DOI:** 10.1101/2025.04.17.25326032

**Authors:** Elyse Letts, Damien Masson, Joyce Obeid

## Abstract

We present iPlayer, a video annotation tool for direct observation of human activity. In contrast to other tools generally designed for continuous or intermittent annotation, iPlayer is optimized for annotations of specific time windows. The tool is cross-platform (MacOS, Windows, and Linux), open-source, locally-run, and configurable to support different labels and time window sizes. Users can annotate multiple activities per window and the video can be optionally looped over the window, reducing annotation error particularly for small window sizes. We developed and used the tool to annotate toddler activity and position in 1-second windows to be synchronized with raw accelerometer data. We also provide examples of how iPlayer could be used with other annotation frameworks. While the tool was optimized for annotations of human activity, we believe it may be of use to any researchers looking to precisely annotate segments of video recordings.

## Introduction

When developing or validating accelerometer methods for human behaviour, having an accurate ground truth is essential (Clevenger et al., 2022; Lyden et al., 2014). A gold-standard for ground truth in human movement analysis is direct observation (Hukka, 2020; Letts et al., 2024; Lyden et al., 2014). A 2024 review found that almost half of the studies that validate methods against direct observation use a video recording (Letts et al., 2024). These video recordings of activities of interest or free-living sessions are then annotated continuously or in fixed windows (Letts et al., 2024). These video annotations can then be linked by time to the accelerometer data and used to train machine learning models or validate existing methods. Key in this process is how the video annotation is completed.

Many tools exist to facilitate this video annotation ranging from manual to fully-automated methods. Automated methods, which generally use video recognition models, can speed up the annotation task by reducing time and personnel requirements, however, they introduce an extra layer of potential bias by using previously developed recognition models (Demrozi et al., 2023). Manual methods, considered the gold-standard, can be very time consuming, so having a tool that facilitates the process is important (Friard & Gamba, 2016).

There are a number of existing video annotation tools that are commonly used. Some tools have a high financial cost (e.g., Noldus Observer XT (Zimmerman et al., 2009)) which may be prohibitive. Others are free, including BORIS (Friard & Gamba, 2016), JWatcher (Stankowich, 2008), and Datavyu (Datavyu Team, 2014). These tools provide comprehensive annotation capabilities supporting many different types of annotations. The flipside of this comprehensive nature is that they may be overly complex for more straightforward video annotation tasks. For example, JWatcher requires at least 3 unique file types to set up the annotations and can generate more than 10 types of output files. Additionally, none of these free tools have the ability to loop over a specific time frame. When very high-resolution annotations are required (e.g., 1 second windows), it can be difficult to annotate on a non-stopping video. Annotators may miss the window, having to constantly go back, or be unclear where a window starts and ends. This could increase the error in the annotations intended to be ground truth, resulting in poorer outcomes if the annotations are used to develop new or validate existing methods. For example, if annotation windows are synchronized with raw accelerometer data windows, having unclear window edges may reduce accuracy of annotations and thus reduce performance of a newly trained machine learning model (Peters et al., 2024).

This brief technical report presents a free, open-source, locally-run tool, named iPlayer. iPlayer is optimized for time-windowed annotations: the video is segmented based on the time window; each segment is looped to help annotators correctly identify the movement happening in the particular segment; and the tool is configurable to support many annotation frameworks. While video annotations tools are not novel, we feel that the tool may be beneficial to other researchers completing similar video annotation tasks and thus are sharing the details of the tool and providing open-source access.

## Description of the tool

iPlayer has three main views, as shown in Figure 1: (A) a video player showing the current video segment being annotated with a few configuration options; (B) a timeline indicating which windows have been annotated and allowing to jump to specific windows; (C) an annotation panel listing all the possible options, as configured by the researchers.

**Figure 1.**
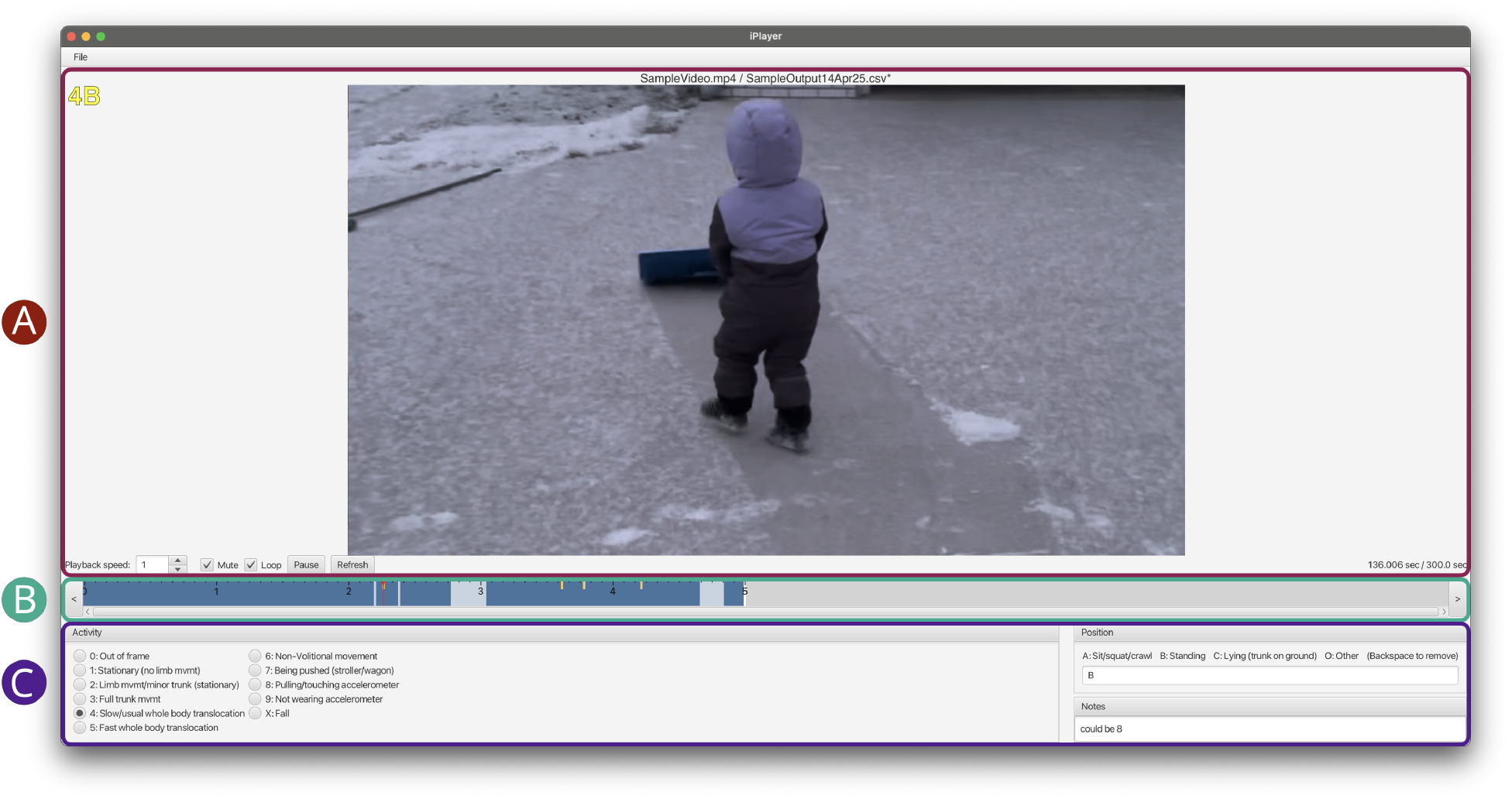
Screenshot of iPlayer in use with three main views highlighted: (A) the video player and its video options (speed, mute, loop, pause/play); (B) the timeline showing windows that are fully-(dark blue), partially-(light blue), or not-(white) annotated with yellow indicating windows with comments and the red showing current position; and (C) the annotation panel showing the annotation framework and Notes section.

The video player area has controls to speed up or slow down the video (playback speed), mute the video, loop the video over the window, and refresh the video player in cases of video freezing. It also shows the annotations (once selected) in yellow for ease of review.

The timeline uses colours to indicate completeness: dark blue indicates that both annotations are complete (e.g., both activity and position), light blue indicates that one annotation is complete (e.g., position but not activity), white indicates that no annotation has been entered, yellow indicates a comment in the Notes section, and the red indicator shows current position in the video. Users can move between time windows by clicking on the timeline, using arrow keys, or using the enter key to advance.

The annotation panel shows the user defined annotation framework. Values can be entered using key shortcuts or by selecting the radio buttons. Notes are entered by clicking into the Notes box which temporarily disables the key shortcuts to allow free typing. Key shortcuts are re-enabled by clicking outside of the Notes box.

Video files are loaded first (File>Load video), followed by either generating a new output .csv file (File>Save as) or by loading a previously started .csv (File>Load csv). The output .csv has the following headings: (1) Index: sequentially numbered windows; (2) Timestamp: the time in seconds from the start of the video for the start of that window; (3) User defined annotation 1 (e.g., Activity in example in Figure 1): the selection from the first annotation; (4) User defined annotation 2 (e.g., Position in example in Figure 1): the selection from the second annotation; (5) Notes: the string entered in the notes section.

### Tool configuration

Labels, shortcuts, and the duration of the time window can be configured in an XML-file placed at the root of the application (the same folder as the application). By modifying this file, researchers can customize the interface to support different annotation tasks. For example, this file replicates the configuration we have been using to annotate toddler data over 1-second time windows using a modified version of the Children’s Activity Rating Scale (Puhl et al., 2013) and adding position labels. The position labels are “multichoice” meaning that more than one label can be entered for a given window. This allows users to capture transitions in position within the time window (e.g., the participant sits up from the floor).

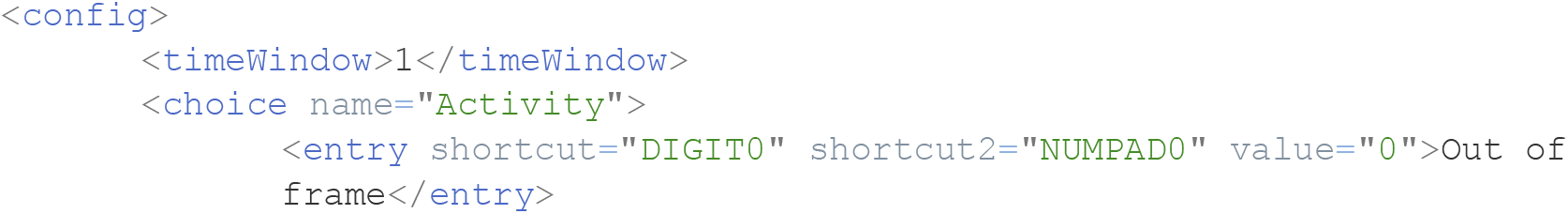
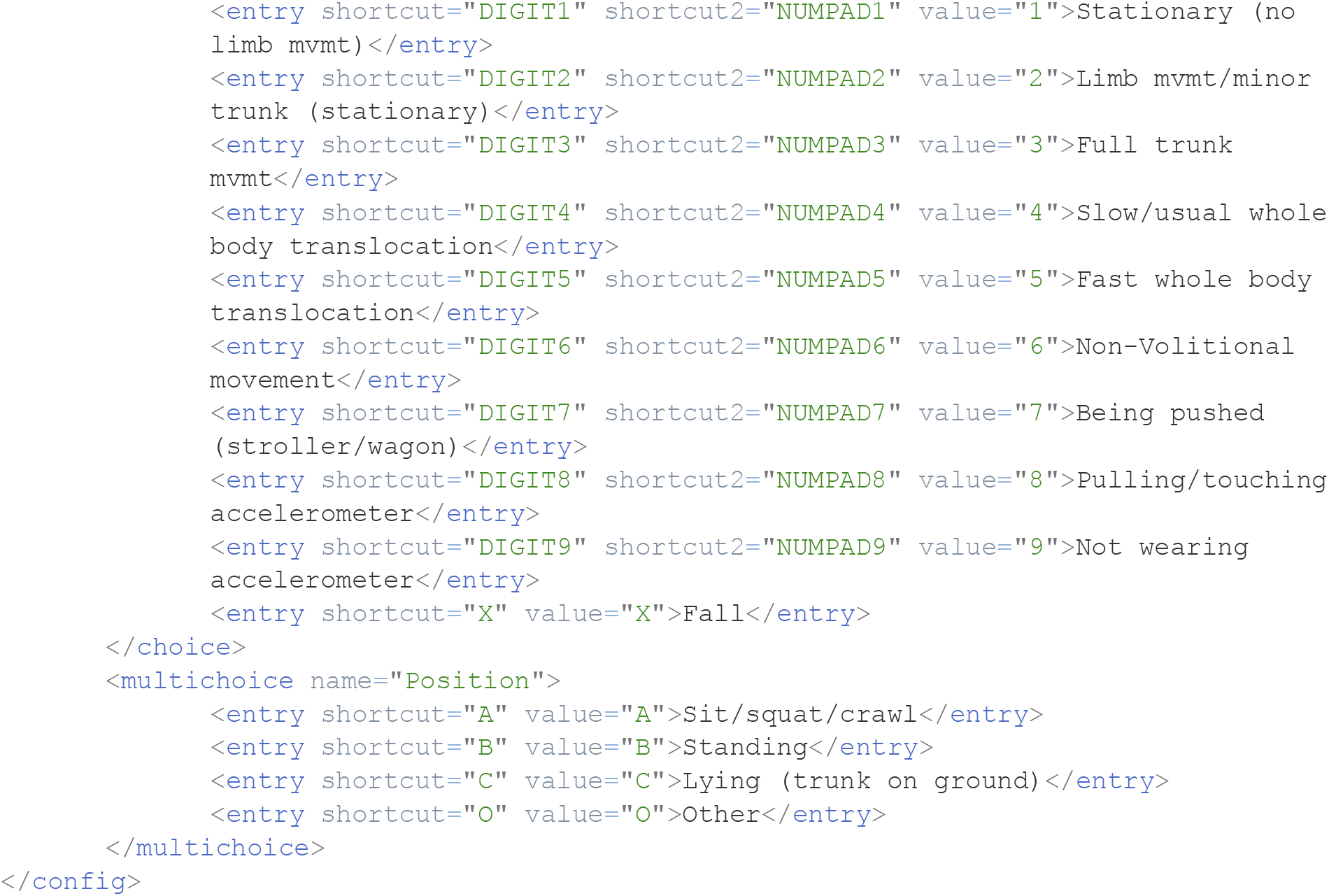

### Implementation

iPlayer was developed in Kotlin (*Kotlin Programming Language*, 2011), using JavaFX (*JavaFX*, 2008) for the graphical user interface. The tool is cross-platform (MacOS, Windows, Linux) and open source. The code and downloadable software application can be found here: https://github.com/LettsE/iPlayer.

## Preliminary Evaluation

We developed iPlayer as a more efficient alternative to Datavyu for our needs to annotate toddler physical activity, sedentary time, and position in one second windows (received ethics clearance from the Hamilton integrated Research Ethics Board; HiREB #3674). The tool was designed through multiple iterations based on the feedback received from annotators. In total, 19 annotators used it (on both Windows and MacOS) to annotate a total of 218 videos. The videos were recordings of toddler play sessions and ranged from 43 to 100 minutes in length.

Annotators had to review and annotate each second of these videos and it took about six hours on average to complete a video. This allowed us to test the robustness of the tool on long videos that were annotated over multiple sessions. While we cannot guarantee the tool works for all users or use cases, we note that all annotators successfully used the tool with no major issue. One annotator commented on how much of an improvement the tool was, specifically the looping feature, over Datavyu for our specific use case.

To further demonstrate the flexibility of iPlayer, we have selected four representative annotation frameworks from the Letts et al. (2024) review and generated their XML configuration files. The four annotation frameworks are: (1) the System for Observing Fitness Instruction Time (SOFIT) as used by Capio et al. (2010) in 15s windows; (2) steps or pedal revolution counts as described by Gatti et al. (2016) in 60s windows; (3) a custom activity list combined with MET classification as used by Hedayatrad et al. (2020) in 5s windows; and (4) a custom activity list designed to match the ActivPAL output including an “other” category as used by Alghaeed et al. (2013) in 1s windows.

For (1) the SOFIT, the XML configuration file could be as follows:

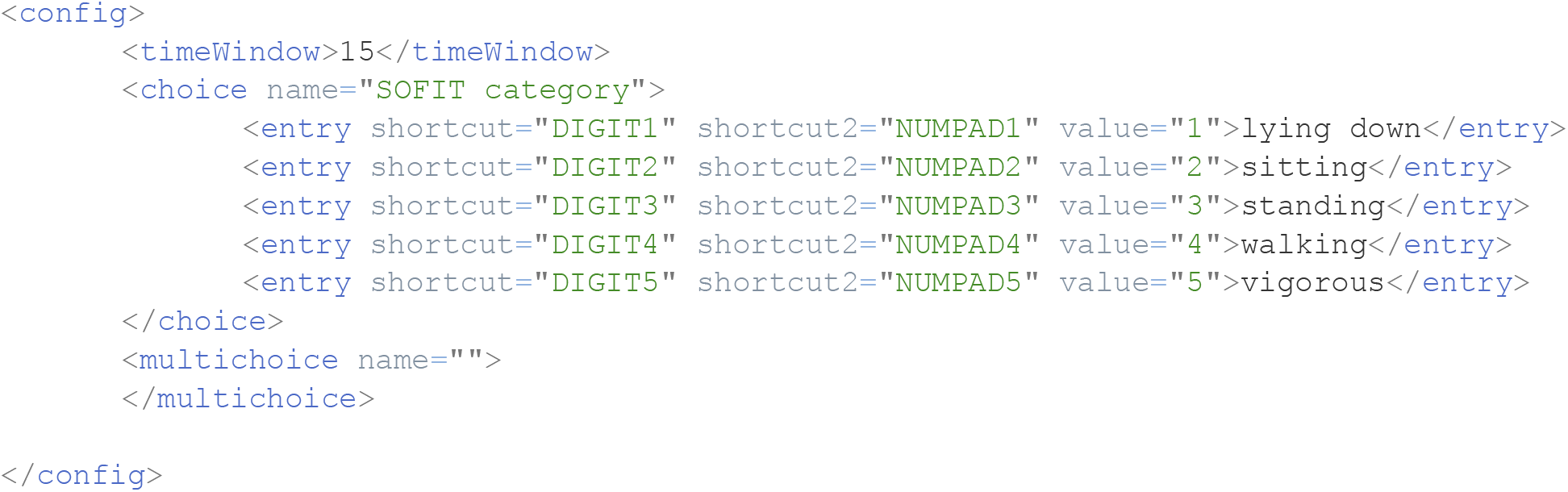

For (2) the step/pedal revolution counting, the configuration file could be as follows with the number of steps/revolutions entered in the notes section:

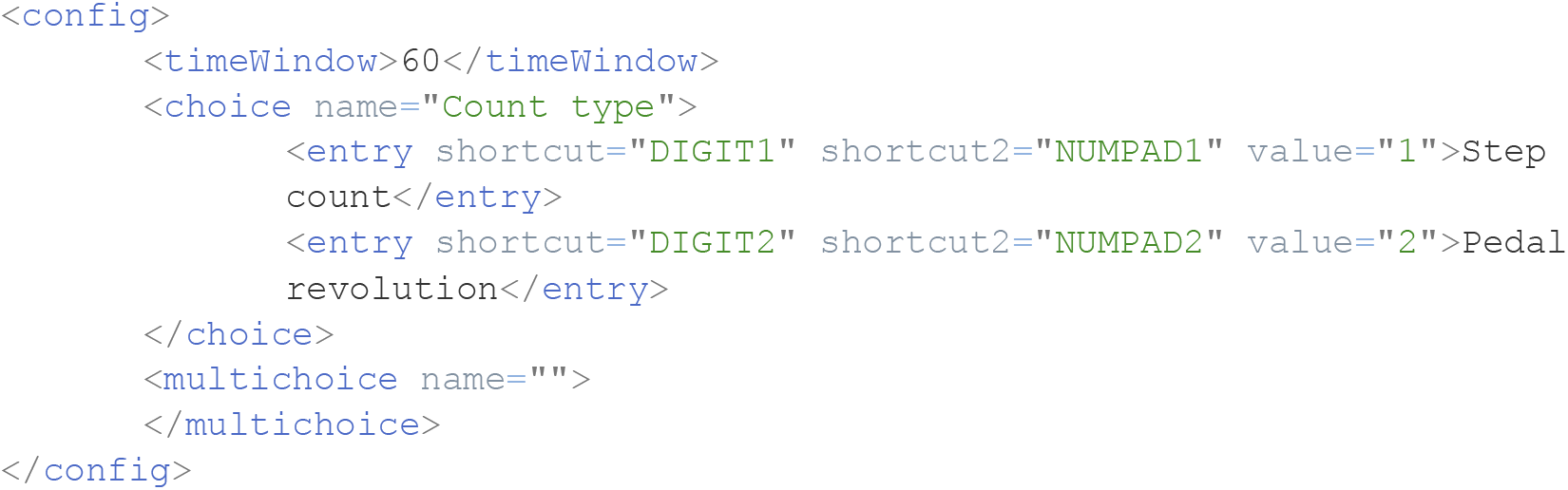

For (3) the custom activity list combined with METs, the configuration file could be:

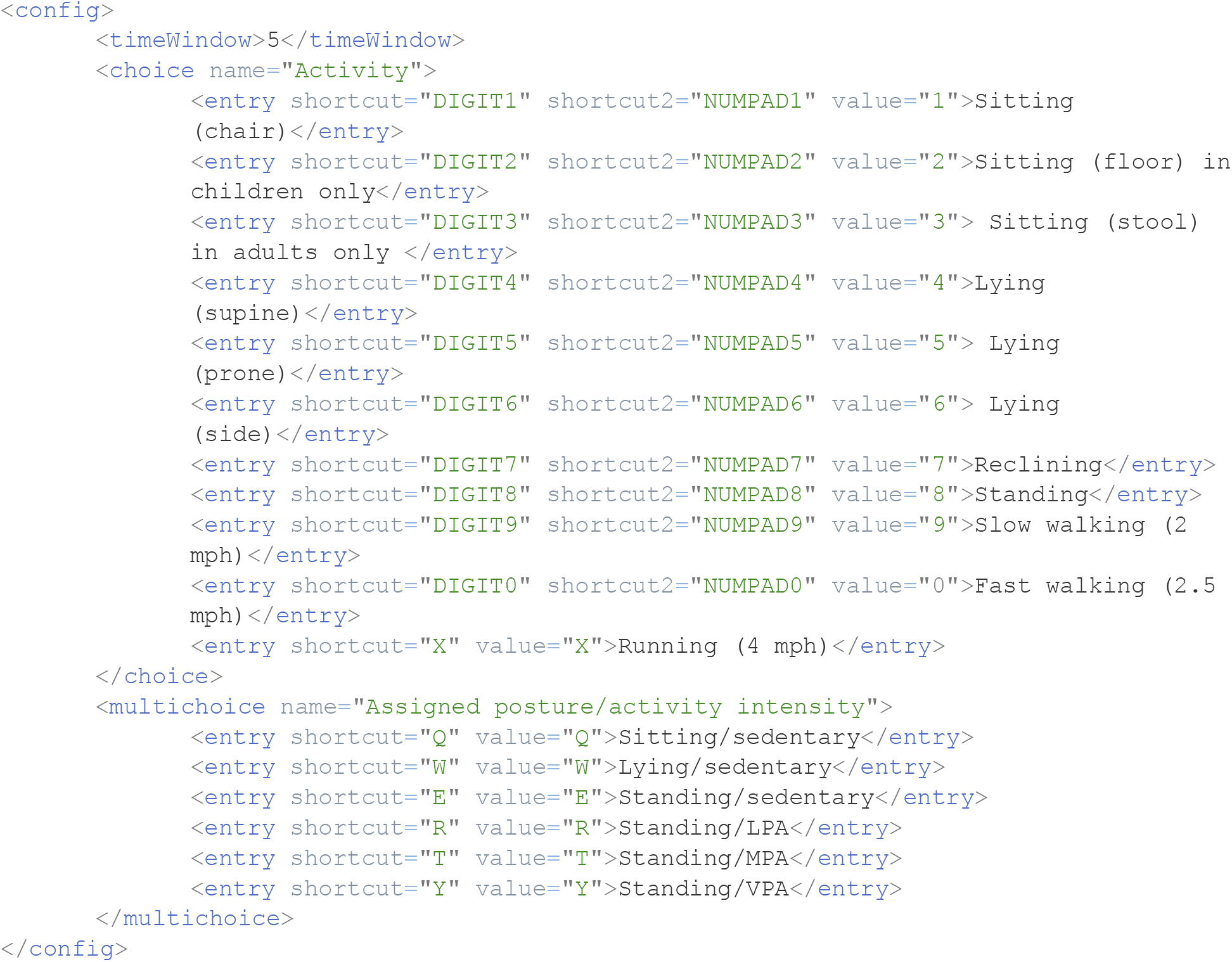

Finally, (4) the custom activity list designed to match the ActivPAL output could have a configuration file as follows, using the Notes section to describe what the “other” is:

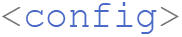
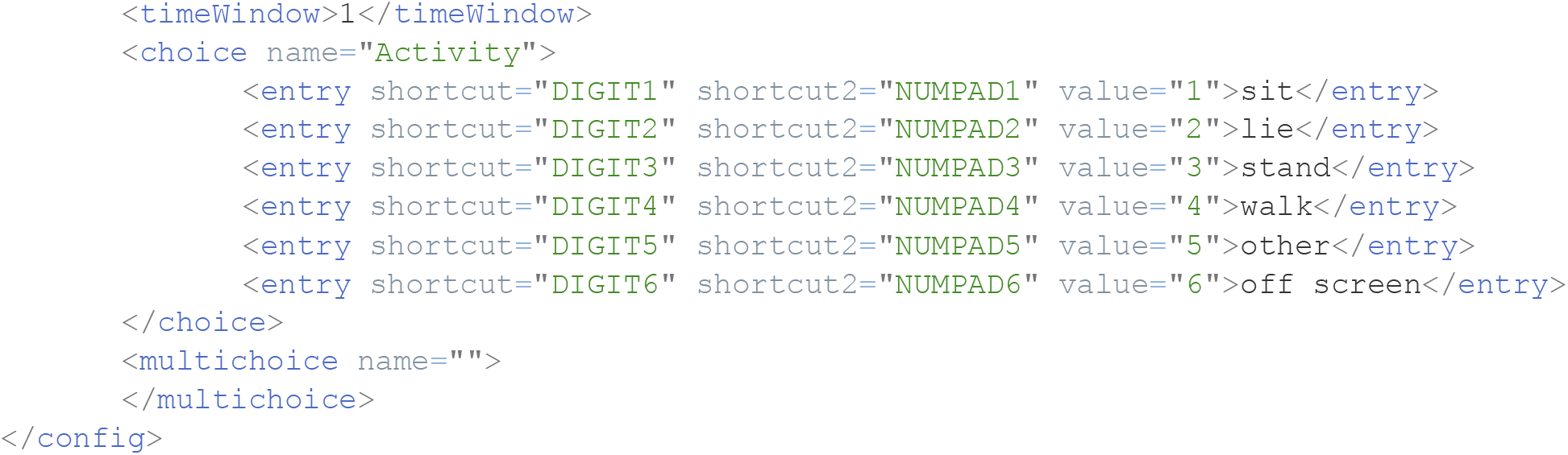

Each of the above sample configuration files can be further customized by users. These examples demonstrate the flexibility of iPlayer to adapt to many annotation frameworks and window sizes.

## Discussion

This paper presents iPlayer, a simple, open-source tool to annotate video recordings in windowed segments. It is designed to help researchers who need to label ground truth activity. It allows for window size and annotations to be customized, with multiple annotations per window as well as a notes feature. It outputs a .csv file which can be saved and reloaded into the tool to revise or resume partially completed annotations. The notes feature can be used to synchronize annotations to other data based on an occurrence in the video (e.g., clapping, clock shown in video).

It may not, however, be suitable for all direct observation needs. iPlayer does not provide a method for live annotation nor for non-windowed (continuous timestamped) annotation. Additionally, users with more complex annotation needs or who need more customization than is currently available may prefer using one of the other existing tools such as BORIS or Datavyu.

## Conclusion

This paper presents iPlayer, an open-source tool to simplify windowed video annotation of human activity and describes an example use cases. It is a video player combined with an annotation tool in which labels and window length can be customized. Its ability to loop the video over a selected window provides a benefit over existing open-source tools and can help improve the accuracy of windowed annotations. We hope that this tool may be of use to other researchers looking to manually annotate video recordings.

## Data Availability

The code and downloadable software application can be found here: https://github.com/LettsE/iPlayer

